# Pushing the Boundaries for Centrally Located Breast Tumors in Oncoplastic Breast Surgery: A Single Centre Audit

**DOI:** 10.1101/2024.11.21.24317412

**Authors:** C.B Koppiker, Aijaz Ul Noor, Sneha Joshi, Rupa Mishra, Priya Sivadasan, Shama Sheikh, Anushree Vartak, Namrata Athavle, Purva Sharma, Mugdha Pai, Chetan Deshmukh, Upendra Dhar, Laleh Busheri, Manasi Munshi

## Abstract

**Background:** Breast Conservative Surgery (BCS) was considered a contraindication for centrally located breast tumor (CLBT), with mastectomy being the preferred treatment. However, advances in oncoplastic techniques now offer promising alternatives by facilitating wider surgical margins and enhancing cosmetic outcomes. This study aims to evaluate surgical, oncological, aesthetic outcome and quality of life (QoL) of oncoplastic breast surgery (OBS) for CLBT.

**Material and Methods:** In this study, 136 patients with CLBT were treated with various oncoplastic techniques including Level I (Grisotti Flap), Level II (Therapeutic Mammoplasty), and Level III (Perforator Flap) techniques. Surgical approach selection was guided by breast-size and degree of ptosis. Oncological and cosmetic outcomes were evaluated by surgeons post-operatively.

**Results:** The mean age of the patient was 52.7 years (range: 22-79). The Level I oncoplasty technique was utilised in 7 patients (11.7%); Level II (Therapeutic Mammoplasty) oncoplasty in 57 patients (41.9%) and Level 3 (Perforator Flaps) in 52 patients (38.2%). 13.9% of post-operative complications were observed. Cosmetic score revealed good to excellent outcomes regarding the patients’ satisfaction toward the surgical procedure. The study observed low rate of local recurrence (4.4%), distant recurrence (5.8%) and metastasis (5.8%). Mortality was 3.6% while all survival (OS) and disease-free survival (DFS) were high at 93.82% and 90.6%, respectively demonstrate the effectiveness of the surgical and adjuvant therapy employed.

**Discussion:** OBS offer viable and effective approach for managing CLBT. These procedures not only achieve satisfactory aesthetic results, but also ensure oncological safety, making them a promising alternate to mastectomy for CLBT.

## Introduction

Oncoplastic breast surgery (OBS), integrating oncologic and plastic surgery principles, is increasingly prevalent, particularly in western nations [1]. The technique involves meticulous planning of excisions, gland reshaping, and repositioning of the nipple areola complex (NAC) for improved aesthetics and symmetry [2]. OBS, including partial mastectomy with breast reduction, is becoming a standard approach for early breast cancer and in selected cases, with locally advanced and large operable breast cancer [3–4].

Centrally located breast tumors (CLBT), are completely or partially located in the projection of the areola up to the chest wall and/or within 2 cm around the areola [5]. CLBT, comprising 5-20% of cases, pose surgical challenges due to higher axillary lymph node metastasis rate, higher possibility of positive margin and invasion of the NAC, lower satisfying cosmetic outcome, and increased local recurrence rate [6–7]. Historically, mastectomy or radical mastectomy was common, due to concerns over local control and multicentricity risk, with conservative surgery contraindicated. However, evolving techniques like oncoplastic breast surgery (OBS) offer alternative choice [8–9]. Despite advancements, selecting optimal surgical approaches for CLBT remains debated, reflecting the complexity of balancing oncologic principles with aesthetic outcomes and local control.

Current studies regarding OBS in CLBT is limited to support the post-surgery outcomes and oncological safety of OBS in CLBT [10–13]. Existing data is characterised by several significant limitations: study design, small sample size, lack of uniform post-surgical radiation therapy, post-surgery outcomes [complications, cosmetic score and patient reported outcome measures (PROMS)] and inadequate follow-up durations. These factors collectively hinder the ability to draw robust conclusions regarding the efficacy and safety of OBS in CLBT patients. Further study is needed to evaluate the oncological outcomes of OBS in CLBT. To address these gaps, further investigation is needed with more robust study designs, including larger sample size, the integration of post-surgery outcomes and radiation therapy where applicable, and extended follow-up durations. Such studies are crucial to accurately evaluate the outcomes, efficacy, and overall benefits of OBS in CLBT patients.

The published data regarding OBS for CLBT remains limited. Hence, we conducted a retrospective study to analyze the clinicopathological profile, post-surgery outcomes, oncological outcome, oncological safety, overall-survival (OS) and disease-free-survival (DFS) in the patients treated with OBS by multidisplinary team management. These findings aim for alternate approach to breast onco-surgeons in integrating OBS techniques into routine clinical practice for improved central breast cancer management.

## Materials and Methods

### Surgical Procedure

At our Breast Unit, we have adopted various surgical procedures for centrally located breast tumors 1) Grisotti Flap Technique, 2) Therapeutic Mammoplasty, 3) Perforator Flaps.

### Surgical Procedure for Grisotti Flap

In this surgical procedure, the nipple–areola complex (NAC) was demarcated first, followed by marking another small circle just below the NAC and also the inframammary sulcus; then, the medial and lateral borders of the flap were drawn extending from the medial and lateral margins of the areolar down to the inframammary fold and converging distally to give a comma-shaped appearance. Then the flap was de-epithelialized, except for circular area close to the defect. An incision was made at the medial and inferior borders of the flap down to the fascia. The flap is undermined from the fascia laterally so it can be rotated upwards and advanced superiorly to fill the defect.

The NAC and tumor were removed during a central quadrantectomy, together with a column of tissue that extended to the pectoral fascia. The borders of the tumor were indicated for intraoperative frozen section investigation. The placement of titanium clips for adjuvant radiation was planned. The medial and inferior margins of the flap were then incised down to the pectoral fascia with wide mobilization of the flap from the pectoral fascia; then, the flap was advanced and rotated to fill the defect with complete suture of the wounds. In selected cases, a second incision was made in the axillary fold to dissect the axillary lymph nodes [14].

### Surgical Procedure for Therapeutic Mammoplasty (TM)

Therapeutic mammoplasty (TM) is an oncoplastic surgical approach that combines oncological safety with aesthetic outcomes for selected breast tumors in moderate-to-large breasts. Following Wise pattern incisions, a central wedge of breast tissue, including the nipple and tumor, is removed. The margin is usually very wide around the tumor. An inferior pedicle is used to carry parenchyma and skin and advanced into the central defect. The nipple is reconstructed in selected cases, hence the need for parenchymal or nipple-areola complex pedicles is avoided in such cases. The pillars are then simply sutured together, and the skin is closed. In selected patients, lateral dissection extends into the axilla for axillary management, including sentinel lymph node biopsy or axillary lymph node dissection as indicated. No separate axillary incision is made [15–17].

### Surgical Procedure for ICAP (MICAP, AICAP, LICAP) and LTAP Flaps

Patients are examined in the standing or sitting position for assessment of tumour location and flap marking. Prior to the surgery, all patients underwent a full clinical history, physical examination, mammography and ultrasound and core biopsy for diagnosis. Staging investigations and treatment decisions were made by a multidisciplinary team. A handheld doppler examination was performed in all patients, pre-operatively and/or intra-operatively, in order to identify the perforators and mark the flap design [18–19].

We used ICAP (MICAP, AICAP, LICAP) and LTAP flaps to reconstruct central quadrant defects [20–21]. The perforator can be located using a unidirectional doppler 1–3 cm and the flap is then designed around the perforator. The long axis of the flap was designed such that the distance between the perforator and the distal portion of the flap was longer than that between the perforator and the furthest part of the defect, in order to facilitate tensionless closure, the flap. The donor sites were primarily closed in all patients.

The patient can be positioned on her back, her side or somewhere between depending upon the size of the flap to be raised. For LICAP/LTAP flaps, the IMF, lateral mammary fold and posterior axillary fold are marked. The perforator vessels are typically located about 1–2 cm posterior to the lateral mammary fold and about 2–3 cm anterior to the posterior axillary fold arising from 3rd to 7th intercostal spaces. The LICAP perforators are usually found in the 4th to 6th intercostal spaces. The LTAP vessels can be traced vertically along the mid-axillary line for about 2–3 cm, in most cases arising from 3rd to 5th intercostal spaces thus differentiating them from the LICAP perforators. The width of the flap is based on the estimated central breast defect and the available donor skin facilitating adequate closure. The length of the flap can be variable and up to 30 cm of the flap can be harvested without vascular compromise and this would again depend up on the amount of tissue needed for the defect. For MICAP flap, the IMF is marked and the medial perforators can be located about 2–3 cm lateral to the sternal edge, about 1–2 cm inferior to the IMF. The flap extends laterally with the base of the flap situated medially. Similarly, the AICAP is based on the perforators located at 6 o’clock position 1–2 cm inferior to the IMF. The flap extends on either sides of the marked perforator and the tissue at 6 o’clock forms the base.

An incision based on the curvature of the breast will usually allow access to the tumour bed and the axilla. After a skin incision was made, we continued the dissection down to the muscle fascia. Once the tumour excision is completed dissection can proceed to identify the perforators. Once seen, the dissection can move to the lateral aspect of the flap and the flap is raised off the back towards the breast and selected perforators.

The flap was elevated in a subfascial manner. Flaps are de-epithelialised as the subdermal plexus contributes to the vascularity of the flap. The dimensions and length of the flap need to be carefully assessed to allow enough reach to the central quadrant defect, while not taking more tissue than is required and allowing for easy donor site closure.

The perforator with visible pulsation was selected as the pedicle perforator. Dissection continues either to the perforators or close to them and stops at a level that allows full mobility of the flap into the required defect. The pedicle was dissected sufficiently deep to obtain a sufficient rotation arc, and the flap was transferred in a propeller fashion. Proper circulation in the flap was verified by capillary refill and a Doppler examination after flap rotation. After the flap was inset, drains were placed beneath the flap. The donor sites were primarily closed in all patients.

### Study Design

In this retrospective, single institutional, observational study, we have collected the medical records of BC patients that have underwent surgeries for centrally located breast tumor at Orchids Breast Health Clinic (OBHC), Pune, during the period of 2013 to 2023. Informed consent was obtained from all patients for collection of study-relevant medical data associated with disease management or routine follow-up visits. Collected data included demography, medical history, clinicopathological characteristics, surgical notes, post-surgery evaluations, chemo-radiation treatment plans, and follow-up details. The details of the study design and patient selection are depicted in **Figure 1-2**.

**Figure 1:**
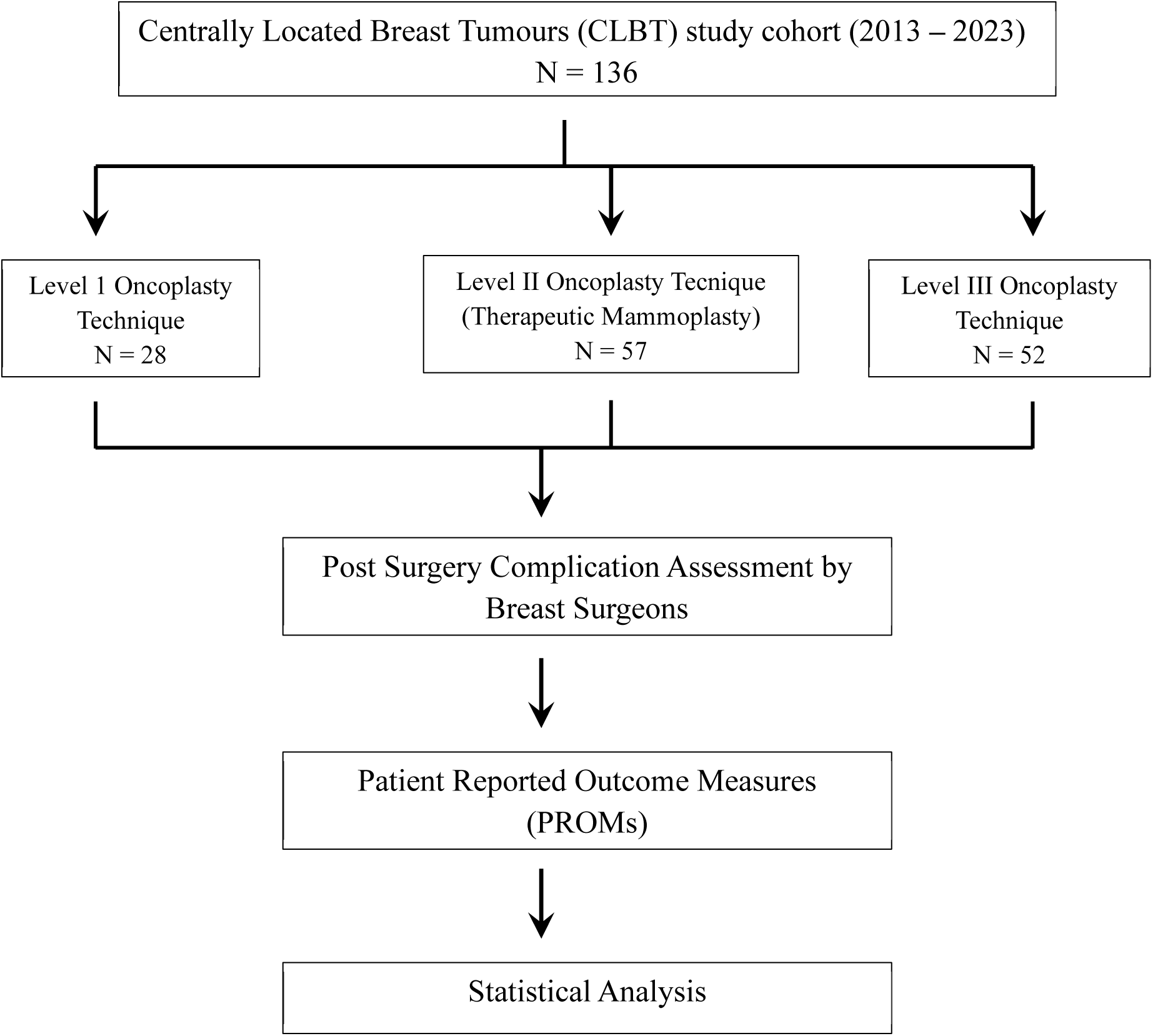
Study flow chart. Distribution of study participants in various groups and assessment of study parameters

**Figure 2:**
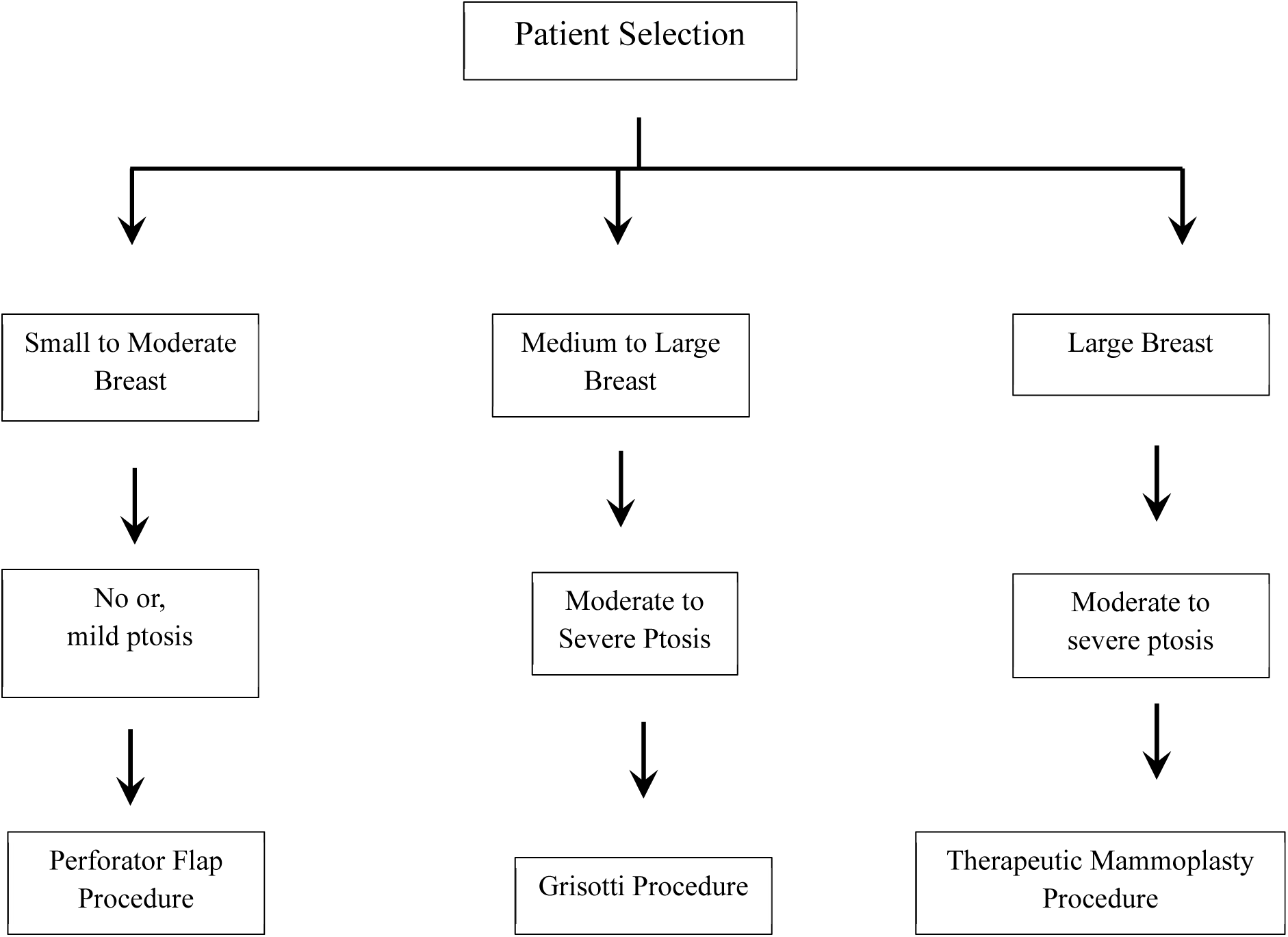
Guide for selection of oncoplastic technique for central quadrant cohort study population.

### Patient Selection

Patient selection at our breast unit is performed by the surgeon to discuss the various treatment and surgical options in a shared decision-making process and detailed pre-op counseling is done. Patients presenting with breast disease in centrally located breast tumors were counseled for viable surgical procedure.

#### Inclusion Criteria For CQ

1) Tumor located in the NAC or located within 2 cm of the edge of the areola

#### Exclusion Criteria For CQ

1) Unknown tumor location,
2) Surgery procedure unknown,
3) Without radiotherapy after OBS,
4) Without follow-up data, or tumor stage
5) Cases whose tumor point of origin could not be determined

### Clinical Management

BC diagnosis was based on clinical examination and radiological evaluation of breast and axilla using full field digital mammography (FFDM) with 3-D tomosynthesis (Siemens Mammomat Inspiration™) and ultrasonography (Siemens Acuson S2000™). Histopathological studies on tru-cut biopsy samples (majority of cases) or vacuum-assisted biopsy (for index tumors, Encor Ultra™) samples were performed for confirming a diagnosis of breast carcinoma. Similarly, ultrasonography and fine needle aspiration cytology were used for investigating axillary lymph node metastasis. Confirmed BC cases underwent surgical procedures for centrally located breast tumors at a network hospital site. The oncologic management with chemo-radiation protocols was undertaken by a multidisciplinary clinical team in accordance with the current NCCN guidelines. Clinical response (clinically complete response (cCR), clinically partial response (cPR), clinically stable disease (cSD), and clinically progressive disease (cPD)) and pathological complete response (pCR) to NACT of the primary tumor were calculated as per Response Evaluation Criteria in Solid Tumors (RECIST) criteria.

### Post-Surgery Outcome Measures

Patient satisfaction and quality of life after IBRS were evaluated using the standardized BREAST-Q questionnaire 12 months post-surgery [22]. The BREAST-Q reconstruction module is divided into multiple independent scales. All scales were scored on a 0- to 100-point scale and higher scores indicated greater satisfaction.

### Statistical Analysis

Data were collected retrospectively from patient records. Follow-up information was taken as recorded in the patient file. The date of recurrence was taken from one of the biopsy pathologies, fine-needle aspiration cytology (FNAC), or PET reports. Overall survival was calculated as all-cause since in many cases it was difficult to ascertain if death was due to disease or other unrelated causes. The overall survival interval was taken as the time period between surgery and death. The exact date of death used in the analysis was in most cases taken as the closest approximation to the date of death as informed by relatives of the patient.

Percent disease-free and percent overall survival were derived from the survival table when the time was the closest median follow-up.Survival rates were analyzed with Kaplan–Meier curves with a log-rank test for the calculation of p-value (<0.05) and Hazard ratio (HR). Survival analysis was plotted using GraphPad Prism 5 for Windows Version 5.01.

## Results

### Central Quadrant Cohort

This is a single institutional audit done for central quadrant tumor for management, tumor response and overall survival. Inclusion criteria for central quadrant tumor located in the nipple areolar complex (NAC) or located within 2 cm of the edge of the areola. All the information was curated from the patient’s file, and follow-up was updated from their last visits or with a telephonic conversation whenever required.

In this study cohort of 171 patients from Central Quadrant cohort undergoing surgery, the distribution of Breast Conservation Surgery (BCS) 80.7% and mastectomy 19.2% was observed. Among the BCS level I oncoplastic technique accounted for 20.6%; level II for 21.6% and level III for 38.2%.

136 patients with centrally located breast tumor diagnosed between 2013 and 2023 who received treatment (NACT/ Surgery/ ACT/RT) at our centre are included in this audit study. Patient demographic details and tumor characteristics for the cohort are summarized in **Table 1**.

**Table 1:** Demographics and tumour characteristics among study patients with oncoplastic surgery (N=138)

| Characteristics | Class Interval | Number | Percentage |
| --- | --- | --- | --- |
| Patient Age (Years) | <=35 | 05 | 3.6% |
|  | 36 - 50 | 60 | 43.4% |
|  | >= 51 | 73 | 52.9% |
|  | Mean Age | 52.08 |  |
| BMI (kg/m<sup>2</sup>) | Mean BMI | 29.89 |  |
| Size of Breast | S | 23 | 16.7% |
|  | M | 61 | 44.2% |
|  | L | 39 | 28.2% |
|  | XL | 01 | 0.7% |
|  | NAV | 14 | 10.14% |
| Ptosis | I | 30 | 21.73% |
|  | II | 46 | 33.4% |
|  | III | 38 | 27.5% |
|  | No | 09 | 6.5% |
|  | NAV | 15 | 10.8% |
| Focality | Unifocal | 101 | 73.1% |
|  | Multifocal | 37 | 26.8% |
| Tumour location | Periareolar | 03 | 2.1% |
|  | Retroareolar | 73 | 52.9% |
|  | Central Quadrant / Deep Central Quadrant | 59 | 42.7% |
|  | NAV | 02 | 1.4% |
|  | NA | 01 | 0.7% |
| Indications of surgery based on Tumour Pathology | IDC | 97 | 70.3% |
|  | ILC | 03 | 2.1% |
|  | IDC + DCIS | 18 | 13.0% |
|  | Others | 15 | 10.9% |
|  | NAV | 05 | 3.6% |
| Tumour receptor status | ER Positive | 86 | 62.3% |
|  | PR Positive | 80 | 58.0% |
|  | ER-PR Positive | 68 | 49.2 |
|  | Her-2 Positive | 29 | 21.0% |
|  | TNBC | 27 | 19.5% |
| Specimen weight (g) |  | Data Not Ava |  |
| Volume of excision (ml) |  | Data Not Ava |  |
| Tumour Size (cm) | < 2 | 36 | 26.0% |
|  | 2-5 | 81 | 58.7% |
|  | >5 | 14 | 10.1% |
|  | NAV | 06 | 4.3% |
| Grade | I | 13 | 9.4% |
|  | II | 91 | 65.9% |
|  | III | 26 | 18.8% |
|  | NAV | 04 | 2.9% |
|  | NA | 04 | 2.9% |
| Stage | 0 | 08 | 5.7% |
|  | I | 36 | 26.0% |
|  | II | 64 | 46.4% |
|  | III | 14 | 10.1% |
|  | NAV | 11 | 8.0% |
|  | NA | 05 | 3.6% |
| Neoadjuvant chemotherapy |  | 49 | 35.5% |
| Adjuvant chemotherapy |  | 65 | 47.1% |
| Adjuvant endocrine therapy |  | 68 | 49.2% |
| Radiation therapy | Yes | 116 | 84.0% |

The cohort’s median age was 52.7 years (range: 22-77), with the maximum number of patients diagnosed in the age group of ≥51 years. The mean body mass index (BMI) was 26.54 (kg/m^2^). Most patients in the cohort had a low-grade tumors (I/II) (77.4%, 93/120) and 23 (19.2%) had high-grade tumors (III). Out of 120 patients, 46 (38.3%) received neo-adjuvant chemotherapy before the primary surgery. After the surgery, 76.7% received radiation-therapy.

### Neoadjuvant Systemic Therapy (NAST)

In this study following NAST, complete response (CR) was seen in 4.3%, partial response (PR) in 54.3% and 21.7% with stable disease. Out of these, pathological complete response (pCR) was achieved in 19.5%. The NAST data was not available for 19.9% of patients. NAST details are summarised in **Table 8**

### Surgery in patients with central quadrant tumors

Data from 136 patients undergoing breast conservation surgery (BCS) in a central quadrant is available and distribution is as follows: Level I oncoplasty in 11.7%; Level II (Therapeutic Mammoplasty; TM) oncoplasty in 41.9% and Level 3 (Perforator Flaps) in 37.5%.

With in Level I, Grisotti Flap, round block and simple oncoplastic closure were employed. The level 2 technique include simple, complex and extreme TM technique. Level 3 include various advanced techniques such-as lateral intercostal artery perforator (LICAP), medial intercostal artery perforator (MICAP), anterior intercostal artery perforator (AICAP) and combinations these of. The most frequent technique in Level 3 was LICAP (14.7%), indicating a preference for advanced oncoplastic techniques. Surgical details are summarised in **Table 2 - 5**.

**Table 2:**
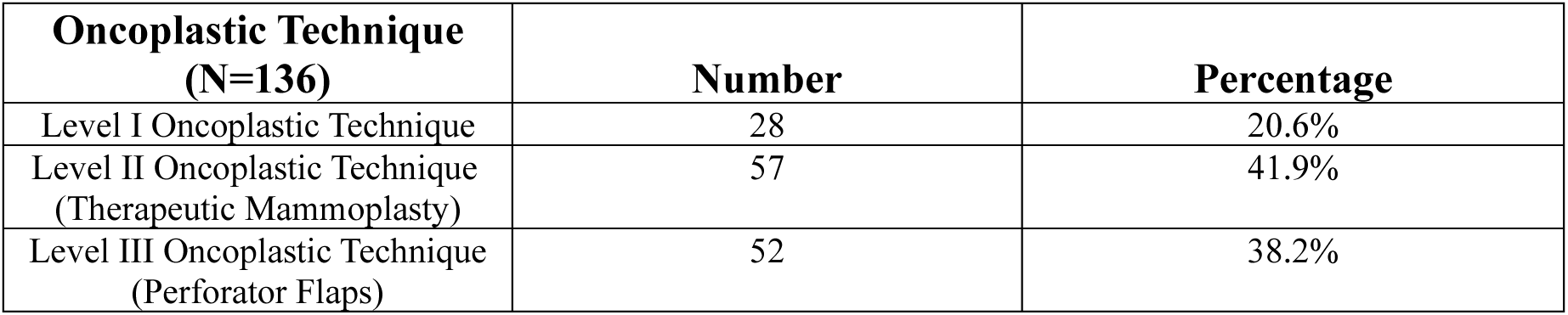
Oncoplastic Technique for Centrally Located Tumor.

| **Oncoplastic Technique**<br>**(N=136)** | **Number** | **Percentage** |
| --- | --- | --- |
| Level I Oncoplastic Technique | 28 | 20.6% |
| Level II Oncoplastic Technique<br>(Therapeutic Mammoplasty) | 57 | 41.9% |
| Level III Oncoplastic Technique<br>(Perforator Flaps) | 52 | 38.2% |

**Table 3:** Types of Level I Oncoplastic Techniques.

| **Level I**<br>**Oncoplastic Technique** | **Number** | **Percentage** |
| --- | --- | --- |
| Grisotti Flap | 07 | 5.1% |
| Round Block | 09 | 6.6% |
| Simple Oncoplastic Closure | 12 | 8.8% |

**Table 4:** Types of Level II Oncoplastic Techniques.

| **Level II**<br>**Oncoplastic Technique** | **Number** | **Percentage** |
| --- | --- | --- |
| Simple Therapeutic Mammoplasty | 28 | 20.5% |
| Complex Therapeutic Mammoplasty | 17 | 12.5% |
| Extreme Therapeutic Mammoplasty | 12 | 8.8% |

**Table 5:** Types of Level III Oncoplastic Techniques.

| **Level III**<br>**Oncoplastic Technique** | **Number** | **Percentage** |
| --- | --- | --- |
| LICAP | 20 | 14.7% |
| AICAP | 09 | 6.6% |
| MICAP | 03 | 2.2% |
| LICAP + AICAP | 01 | 0.7% |
| LTAP | 09 | 6.6% |
| LTAP + LICAP | 10 | 7.3% |

### Post-Operative Complications

Post-operative complications were classified based on grades as per the Clavien–Dindo classification adapted for breast cancer. In this cohort, a total of 13.9% of complications were observed. **(Table 6)** All complications were treated conservatively in the outpatient setting. In general, we observe immediately post-surgery a low rate of Grade III complications even with level II and level III techniques.

**Table 6:** Post-op complications in the cohort as per the Clavien – Dindo Classification.

| Characteristics | Complications |  |  |
| --- | --- | --- | --- |
|  | Level I<br>Oncoplastic<br>Technique | Level II<br>Oncoplastic Technique<br>(Therapeutic Mammoplasty) | Level III<br>Oncoplastic<br>Technique<br>(Perforator Flaps) |
| Grade I<br>(Seroma/Hematoma not requiring<br>drainage; minor skin-necrosis; fat<br>necrosis; delayed wound healing) | 01 | 06 | 2 |
| Grade II<br>(wound infection) | – | 01 | 1 |
| Grade IIIa<br>(Seroma/Hematoma were drained<br>under USG guidance; lymphedema;<br>nipple necrosis; skin-necrosis<br>undergoing debridement) | – | 01 | - |
| Grade IIIb<br>(seroma/hematoma drained under<br>general anesthesia—major skin<br>necrosis, wound infection requiring<br>debridement, bleeding) | – | 01 | 06 |
| Total | 01 | 09 | 09 |

### Cosmetic Score

Out of 120 surgeries for centrally located breast cancer patients, cosmetic scores were assessed by surgeons within 3–6 months post-surgery. The evaluation criteria were the form and symmetry of the breast and satisfaction of the patient. The post-operative cosmetic results evaluated by the surgeon was good in 66.1% cases and excellent in 2.9% cases. **Table 7** shows the cosmetic scores as reported by the surgeons.

**Table 7:** Cosmetic score for surgeries in Breast Cancer patients.

| Category | Score | Classification | Level I<br>Oncoplastic<br>Technique | Level II<br>Oncoplastic<br>Technique<br>(Therapeutic<br>Mammoplasty) | Level III<br>Oncoplastic<br>Technique<br>(Perforator<br>Flaps) | Number | %age |
| --- | --- | --- | --- | --- | --- | --- | --- |
| 1 | 0-3 | Bad | - | - | - | - | - |
| 2 | 4-5 | Fair | 02 | 02 | 04 | 08 | 5.8% |
| 3 | 6-8 | Good | 13 | 48 | 29 | 90 | 66.1% |
| 4 | 9-10 | Excellent | 01 | 03 | - | 04 | 2.9% |
|  |  | Total | 16 | 53 | 33 | 102 | 74.8% |
| 5 | NA | Not<br>available | 12 | 04 | 19 | 35 | 25.7% |

**Table 8:**
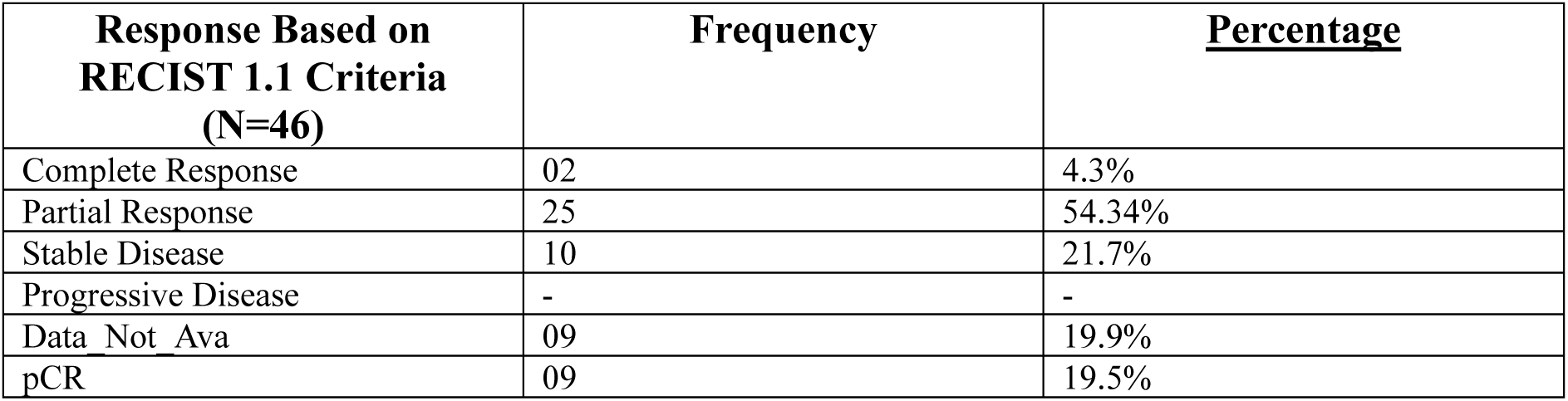
Response based on RECIST 1.1 in the central quadrant study population.

| Response Based on<br>RECIST 1.1 Criteria<br>(N=46) | Frequency | <u>Percentage</u> |
| --- | --- | --- |
| Complete Response | 02 | 4.3% |
| Partial Response | 25 | 54.34% |
| Stable Disease | 10 | 21.7% |
| Progressive Disease | - | - |
| Data Not Ava | 09 | 19.9% |
| pCR | 09 | 19.5% |

### Patient-reported outcome measures (PROMs)

PROM data were collected from the study participants after a minimum period of 12 months post-surgery using the BREAST-Q questionnaires. Out of 136 breast cancer patients, 109 (86%) responded to the questionnaire. High patient satisfaction scores were observed from our PROM data as seen in **Figure 5**.

### Oncology and Survival Outcomes

The median follow-up for the study was 35.5 months. The study observed low rate of local recurrence (4.4%), distant recurrence (5.8%) and metastasis (5.8%). **(Table 11)** Mortality was 3.6% whereas overall survival (OS) and disease-free-survival (DFS) were high at 93.82% and 90.6% respectively as seen in **Figure 3 and 4**.

**Figure 3:**
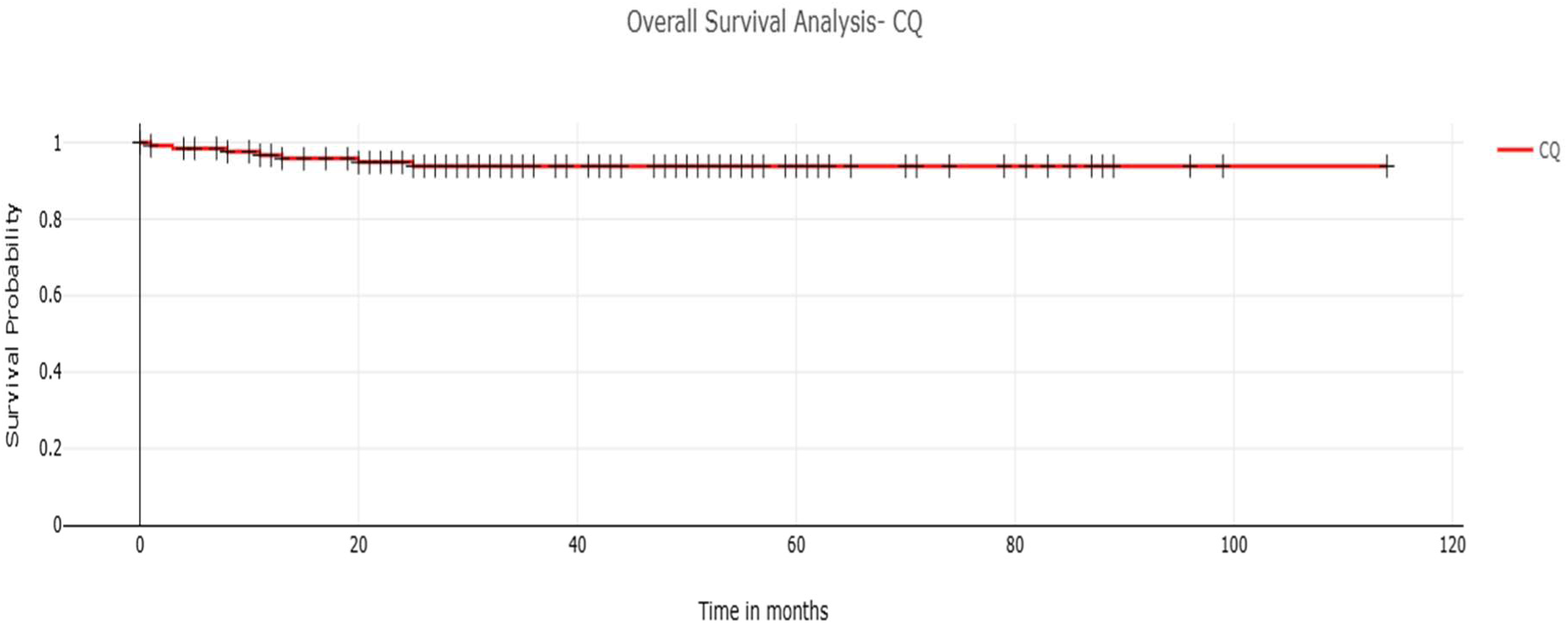
Overall Survival Analysis.

**Figure 4:**
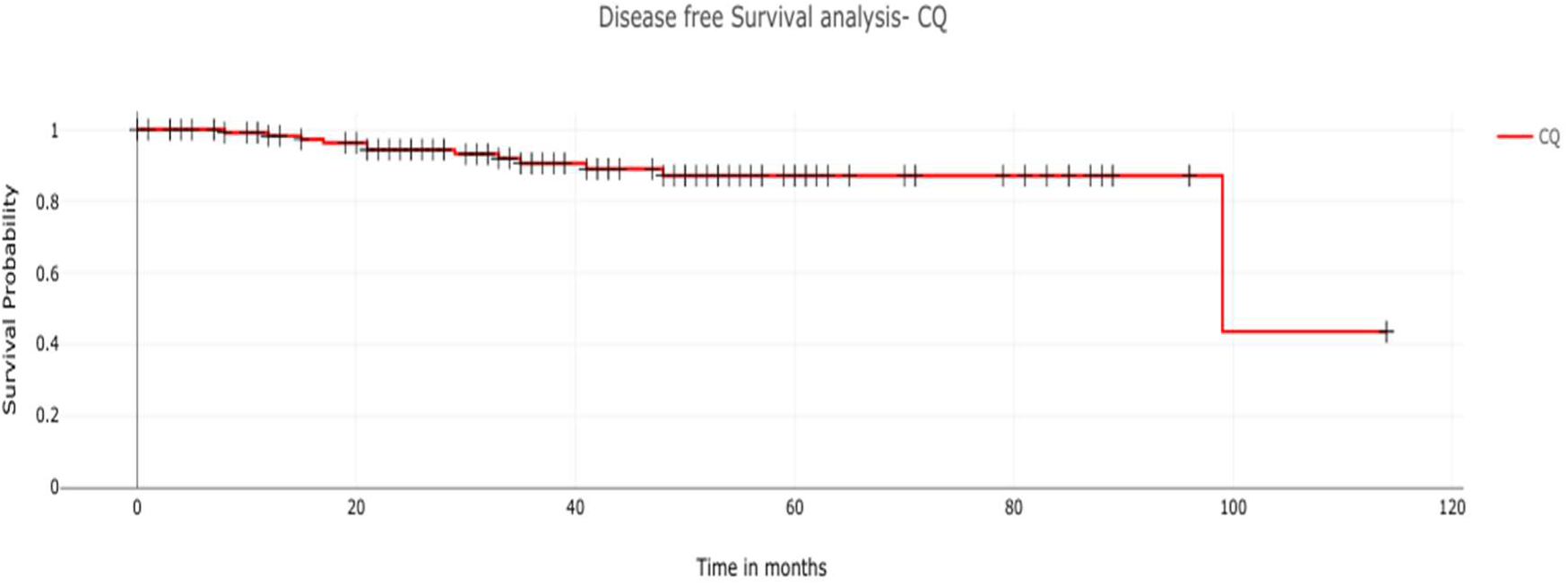
Disease Free Survival Analysis.

**Figure 5:**
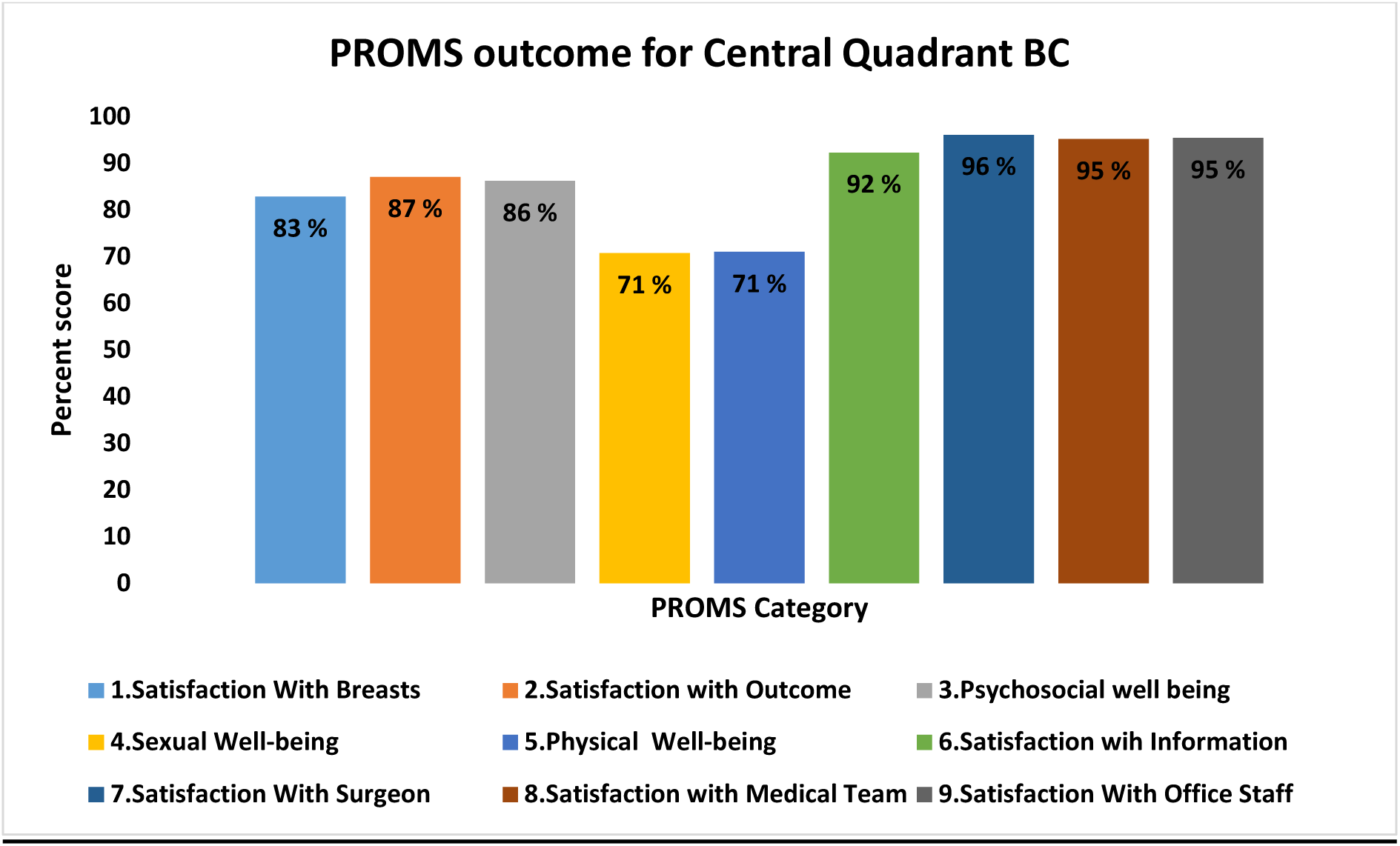
Analysis of PROMS.

### Triple Negative Breast Cancer (TNBC) Cohort

TNBC cohort exhibited a mean age of 41.8 years. Tumor grading revealed that significant portion had a high-grade tumor (Grade III), comprising 44.5% of the cohort, while 37.0% had a low-grade tumor. Tumor staging indicated that the majority of the patient had a T2 tumor accounting for 55.6% of cases. In terms of disease stage at diagnosis, 81.5% were identified at an early stage (<IIB), whereas 19.5% presented with a late stage (>IIB).

Following NAST for TNBC, the clinical response was categorised as follows: 3.7% of patients achieved a CR, 29.6% experienced a partial response and 3.7% had a stable disease. Notably, a pCR was attained by 7.4% of patients. Additionally, NAST data was unavailable for 14.8% of cohort. **(Table 10)**

Regarding treatment modalities, 46.8% underwent NAST prior to primary surgery. Post-surgery, a substantial 84.6% of patients received RT. These demographic and treatment details are comprehensively summarised in **Table 9**.

**Table 9:** Demographic table for central quadrant TNBC study population.

| Characteristics | TNBC Cohort<br>(N=27) | %age | NACT Cohort<br>(N=13) | %age |
| --- | --- | --- | --- | --- |
| **Age (Years)** |  |  |  |  |
| <=35 | 02 | 7.4% | - | - |
| 36-50 | 14 | 51.8% | 08 | 61.5% |
| >=51 | 11 | 40.7% | 05 | 38.0% |
| Mean Age | 41.8 |  | 46.8% |  |
| **Menopause Status** |  |  |  |  |
| Premenopausal | 09 | 33.4% | 04 | 53.8% |
| Postmenopausal | 15 | 55.6% | 04 | 30.7% |
| Not Available | 03 | 11.0% | 02 | 15.0% |
| **Tumor Size** |  |  |  |  |
| T0 | 03 | 11.2% | 02 | 15.3% |
| T1 | 07 | 26.0% | 03 | 23.0% |
| T2 | 15 | 55.6% | 05 | 38.4% |
| T3 | 02 | 7.4% | 03 | 23.0% |
| **Node Status** |  |  |  |  |
| Node Positive | 10 | 37.0% | 05 | 38.4% |
| Node Negative | 17 | 63.0% | 07 | 53.8% |
| Not Available | - | - | 01 | 7.6% |
| **Tumor Grade** |  |  |  |  |
| Low (I/II) | 10 | 37.0% | 05 | 38.4% |
| High (III) | 12 | 44.5% | 05 | 38.4% |
| Not Available | 05 | 18.5% | 03 | 23.0% |
| **Tumor Stage** |  |  |  |  |
| Early (<IIB) | 22 | 81.5% | 10 | 77.0% |
| Late (>IIB) | 04 | 14.9% | 03 | 23.0% |
| Not Available | 01 | 3.7% | - | - |
| **Surgery** |  |  |  |  |
| Level I | 02 | 7.4% | 02 | 15.4% |
| Level II | 14 | 51.8% | 05 | 38.5% |
| Level III | 10 | 37.0% | 06 | 46.1% |
| Not Available | 01 | 3.7% | - | - |
| **Radiation Therapy** |  |  |  |  |
| Radiation Therapy | 24 | 88.9% | 11 | 84.6% |
| Not Available | 03 | 11.1% | 02 | 15.4% |
| **Oncology Outcome** |  |  |  |  |
| Local Recurrence | 03 | 11.0% | 01 | 7.6% |
| Distant Recurrence | 02 | 7.4% | 01 | 7.6% |
| Metastasis | 02 | 7.4% | 02 | 15.3% |
| Mortality | 03 | 11.0% | 03 | 23.0% |

**Table 10:** Response based on RECIST 1.1 for central quadrant TNBC study population.

| Response Based on<br>RECIST 1.1 Criteria<br>(N=27) | Frequency | Percentage |
| --- | --- | --- |
| Complete Response | 01 | 3.7% |
| Partial Response | 08 | 29.6% |
| Stable Response | 01 | 3.7% |
| Progressive Disease | - | - |
| Not Available | 04 | 14.8% |
| pCR | 02 | 7.4% |

**Table 11:** Oncology outcome for Central Quadrant Cohort.

| **Follow-Up Status** | **Number** | **Percentage** |
| --- | --- | --- |
| Local Recurrence | 06 | 4.4% |
| Distant Recurrence | 08 | 5.8% |
| Metastasis | 08 | 5.8% |
| Mortality | 05 | 3.6% |

## Discussion

The comprehensive analysis of CLBT cohort undergoing surgery primarily in the central quadrant provides significant insights into current practices, outcomes, and challenges associated with OBS. The finding underscores the evolving trends and efficacy of oncoplastic techniques in managing the central quadrant tumors, highlighting their implications for management of the patients and outcomes.

Central breast tumors, have been denied the opportunity of breast conservation, given the possibility of multifocality, multicentricity, direct invasion of the nipple–areolar complex (NAC), and aesthetic revulsion arising from the possible removal of the NAC [23]. Clinical trials have proved that breast conservative surgery (BCS) in centrally located tumor is similar to those who undergo mastectomy in terms of local recurrence, disease-free, or overall survival rates [24–25]. However, the conventional conservative treatment or central quadrantectomies, which includes excision of the NAC and the correspondent underlying cylinder of parenchyma down to the pectoralis fascia, may result in local glandular defects and poor aesthetic outcome including obvious distortion of breast contour and scar contracture in most cases [26]. The failure of classical BCS techniques to offer solutions for challenging scenarios has stimulated the advancement of oncoplastic surgery to allow wide excision for BCS without compromising the natural shape of the breast [27]. Several oncoplastic techniques can be used to reconstruct the breast after central quadrantectomy. The choice of the oncoplastic technique depends on tumor size, NAC involvement, the breast volume, and its ptotic degree [28].

In the CLBT cohort, OBS was the predominant surgical approach, utilized in 80.7% of patients, while 19.2% underwent mastectomy. This study showed that OBS offer an excellent option for breast reconstruction in women with small to large-sized breasts, with high patient-reported outcome measures, good aesthetic outcomes, minimal complications and prominence of Level II and Level III oncoplasty techniques. The most important finding of this study is low rate of local and distant recurrence with high OS and DFS.

### CLBT Cohort

In our study, the mean age is 52.8 years. IDC was the most common pathological subtype of breast cancer in this study (70.3%). Similar results have been documented by *Gardfjel et al., Farouk et al., and Park et al.,* which showed an incidence of IDC 78.4, 68.5%, and 80%, respectively [29–31]. The patient’s cosmetic outcome was evaluated as good in 90 patients (66.1 %) and excellent in 04 patients (2.9%). Our results are similar to those reported by Moustafa et al. being good in 55% of his patients [32].

### Oncoplastic Surgery in CLBT

Among the OBS procedures, a diverse range of oncoplastic techniques were employed: Level I Oncoplasty accounted for 20.6%, Level II for 21.6% and Level III for 38.2%. This distribution reflects a clear preference for advanced oncoplastic techniques, Level III being most utilised.

The Level III Oncoplasty which includes complex reconstructive technique such as perforator flaps demonstrates a trend towards patient-centred approaches. The high use of this techniques indicate a commitment to achieve optimal cosmetic outcomes and addressing the challenge associated with central quadrant resections, while preserving breast aesthetics can be particularly challenging. *Nigam et al.* have used CWPFs to reconstruct defects in central quadrant tumours. They report no wound complications and very good-excellent aesthetic outcomes the patients [33]. Similarly, in our study, we observed minimal post-op complications and good cosmetic outcome with CWPFs.

The Level II oncoplasty (Therapeutic Mammoplasty) was also significantly utilised (41.9%). This approach strikes between oncological safety and aesthetic outcomes, reflecting its role in effectively managing larger resections while maintaining a favorable cosmetic result. However, the lower usage of Level I oncoplasty technique (20.6%) suggests that simpler techniques are less favored in the context of more complex cases where advanced oncoplastic methods offer better outcomes.

In our institute, patients with CLBT were managed as follows: whenever there is a great suspicion of NAC involvement preoperatively (suspicious findings within 2cm of the nipple proved by radiological assessment) in patients central quadrantectomy was done. Overall, seven cases (5.1%) were subjected to Grisotti technique, fifty-seven (41.9%) for therapeutic mammoplasty and fifty-two (37.5%) underwent perforator flaps oncoplastic surgery. When lesions were found at a sufficient distance from the NAC in small to moderate breasts, perforator flaps (twenty-six cases) were done. However, in larger breasts, therapeutic mammoplasty was the optimal technique; fifty-seven cases were subjected to this technique. Grisotti technique was the best option for moderate to large breast; seven cases were done in our institute. Similar to our study, McCulley et al., have also used the central quadrantectomy technique that largely depends on the tumor-to-breast size ratio, the degree of ptosis, and the preference of the surgeon, through central elliptical excision of the tumor with direct closure of the defect [34]. However, Grisotti technique has become more popularly used, especially when there is adequate breast size or ptosis with the distance between the nipple and the inframammary fold being at least 7-8cm.

### NAST and Response

The NAST data highlights the impact of NAST on treatment outcome, with 35.5% of patients receiving this therapy prior to surgery. The response to NAST was varied, with 4.3% of CR, 54.3% of PR and 19.5% of pCR. These results indicate that NAST can reduce the tumor burden and improve surgical outcomes underscoring the need for ongoing advancements in systemic therapy.

### Post-Operative Outcomes and Complications

Post-operative complications were relatively low, at 13.9%, and managed conservatively. Notably, Grade III complications were rare even with advanced oncoplastic techniques (Level II and Level III). This suggests that while advanced techniques are more complex, they do not necessarily increase the risk of severe complications, like due to improved surgical precision and post-operative care.

Cosmetic outcomes were favorable, with 6.1% of patients rated ad having good results and 2.9% excellent outcomes. High patient satisfaction scores from PROMs further reinforce the positive impact of oncoplastic techniques on aesthetic outcomes, which is critical for patient quality of life (QoL).

### Oncology and Survival Outcomes

The cohort’s 35.5-month median follow-up revealed promising oncological outcomes: low rates of local recurrence (4.4%), distant recurrence (5.8%), and metastasis (5.8%). High overall survival (OS) and disease-free survival (DFS) rates (93.82% and 90.6%, respectively) demonstrate the effectiveness of the surgical and adjuvant therapy employed. These outcomes reflect the success of combining advanced surgical techniques with effective systemic treatment.

### TNBC Presentation

In the TNBC cohort, the mean age was younger (41.8) years, and a higher proportion had high grade tumors (44.5%). Despite the aggressive nature of TNBC, early-stage diagnosis was prevalent (81.5%), which is favorable for treatment outcomes. NAST yielded lower pCR (7.4%) compared to the overall cohort, which may reflect the distinct biological behavior of TNBC. Post-surgery, a significant proportion received radiation therapy (84.6%, which is consistent with the aggressive nature of TNBC and the need for comprehensive local control.

### Limitations of the study

This study has some potential limitations. The sample size of the CLBT is small. The further research with large cohorts, rigrous study design and comprehensive follow-up is necessary to substantiate these initial promising clinical and PROM observations. Furthermore, this is a single institutional study in which all CLBT procedures are performed by the same operating team. In the future, this study design needs to be replicated in a multi-centric setting in order to validate the findings by avoiding investigator bias. In our cohort no advanced cosmetic procedures that could potentially enhance aesthetic outcomes were performed. This approach underscores the impact of resource limitations on the scope of available treatment in resource-limited healthcare settings.

## Conclusion

This comprehensive analysis underscores the significant advancements in breast oncoplasty, demonstrating a shift towards advanced techniques (Level II and Level III) to balance oncological efficacy and cosmetic results. The favorable outcomes, low complication rates and high patient satisfaction emphasize the benefit of OBS. However, variation in response to NAST and the aggressive nature of TNBC highlight the need for continued research and individual treatment strategies. Overall, the data supports the continued evolution and refinement of oncoplastic techniques to enhance patient outcome and satisfaction in CLBT.

## Data Availability

All data produced in the present study are available upon reasonable request to the authors

## Acknowledgement

We would like to acknowledge unstinting support of Research Grant from BAJAJ AUTO CSR (GC2528). We would also like to thank IRMI -DBT/WT India Alliance fellowship received by Dr. Sneha Joshi. We would also thank clinic staff at Orchids Breast Health and research staff of Centre for Translational Cancer Research for being of immense help at each step. We would like to thank patients consented to be part of the study.

---

**Supplementary Table 1:** Demographics and tumour characteristics among study patients with mastectomy (N=33)

| Characteristics | Class Interval | Number | Percentage |
| --- | --- | --- | --- |
| Patient Age (Years) | <=35 | 06 | 18.18% |
|  | 36 - 50 | 16 | 48.5% |
|  | >= 51 | 11 | 33.34% |
|  | Mean Age | 41.27 |  |
| BMI (kg/m<sup>2</sup>) |  | Data Not Ava |  |
| Size of Breast | S | 07 | 21.2% |
|  | M | 10 | 30.3% |
|  | L | 07 | 21.2% |
|  | XL | - | - |
|  | NAV | 09 | 27.2% |
| Ptosis | I | Data Not Ava |  |
|  | II | Data Not Ava |  |
|  | III | Data Not Ava |  |
|  | No | Data Not Ava |  |
| Focality | Unifocal | 22 | 66.7% |
|  | Multifocal | 11 | 33.3% |
| Tumour location | Periareolar | - | - |
|  | Retroareolar | 07 | 21.2% |
|  | Deep Central Quadrant | 26 | 78.8% |
| Indications of surgery based on Tumour Pathology | IDC | 21 | 63.6% |
|  | ILC | 01 | 03.0% |
|  | IDC + DCIS | 02 | 06.0% |
|  | Others | 06 | 18.18% |
|  | NAV | 03 | 09.10% |
| Tumour receptor status | ER Positive | 18 | 54.5% |
|  | PR Positive | 16 | 48.5% |
|  | ER-PR Positive | 16 | 48.5% |
|  | Her-2 Positive | 05 | 09.1% |
| Specimen weight (g) |  | Data Not Ava |  |
| Volume of excision (ml) |  | Data Not Ava |  |
| Tumour Size (cm) | < 2 | 07 | 21.2% |
|  | 2-5 | 16 | 45.5% |
|  | >5 | 06 | 18.2% |
|  | NAV | 05 | 15.1% |
| Grade | I | 02 | 6.06% |
|  | II | 17 | 51.5% |
|  | III | 07 | 21.2% |
|  | NAV | 06 | 18.2% |
|  | NAV |  |  |
| Neoadjuvant chemotherapy | Yes | 14 | 42.4% |
| Adjuvant chemotherapy | Yes | 19 | 57.6% |
| Adjuvant endocrine therapy | Yes | 12 | 36.4% |
| Radiation therapy | Yes | 10 | 30.3% |
|  | No | 11 | 33.4% |
|  | NAV | 12 | 36.3% |

